# Navigating Life After Dementia Diagnosis: Questions, Challenges, and Support Needs Among Dementia Dyads

**DOI:** 10.64898/2026.10.07.26364992

**Authors:** Katherine C. Britt, Alaa Harb, Brianna Bailey, Bridget Saffold, Xiao Luo, Rebecca T. Brown, Bin Huang

## Abstract

**Background:** Older adults living with mild cognitive impairment (MCI) or Alzheimer’s disease and related dementias (AD/ADRD) and their care partners face substantial daily challenges that evolve with disease progression and extend far beyond the clinical setting, including symptom management, care coordination, healthcare navigation, and planning for future care needs. Understanding these challenges is essential for developing innovative solutions to provide ongoing and targeted support. In this study, we explored questions, challenges, and support needs dementia dyads experienced following diagnosis to inform the development of an artificial intelligence (AI)-based chatbot designed to provide ongoing support for dementia dyads.

**Methods:** We recruited 11 community-dwelling dyads comprising persons living with MCI or mild AD/ADRD and their care partners, one additional care partner, and 12 dementia care experts from academic medical centers, clinics, and aging organizations in the U.S. We conducted semi-structured interviews and analyzed them using conventional content analysis.

**Results:** Participants described interconnected questions, challenges, and support needs that evolved while navigating life with MCI or AD/ADRD. Questions centered around day-to-day care management, medical concerns, behavioral symptom management, diagnosis and prognosis, safety, and resources and services. Challenges included care partner overwhelm, limited dementia knowledge, condition-related uncertainty, healthcare system navigation difficulties, financial constraints, and technology-related barriers. Participants identified needs for ongoing education, personalized care planning, resource navigation, logistical support and follow-up, culturally responsive care, and social and emotional support.

**Conclusions:** Dementia dyads experience substantial informational, emotional, and practical support needs in the early stages of MCI and AD/ADRD. Findings suggest that post-diagnostic support should extend beyond information provision alone to provide ongoing, personalized, and practical guidance that helps dyads navigate changing needs, resources, and everyday dementia care. These findings are informing the development of an AI-based chatbot intended to complement existing dementia care by extending access to education, practical guidance, and resource navigation between clinical encounters.

## Background

Alzheimer’s disease and related dementias (AD/ADRD) affect millions of older adults worldwide and represent one of the leading causes of disability, dependency, and healthcare utilization among aging populations.(*ADI - Dementia Statistics*, n.d.). Although disease-modifying therapies remain limited, early diagnosis of mild cognitive impairment (MCI) and early-stage dementia provides an important opportunity to optimize symptom management, support care planning, promote safety, and improve quality of life for both persons living with dementia and their care partners (Fowler et al., 2025; Frederiksen et al., 2024). However, dementia is a progressive condition characterized by continually evolving cognitive, functional, behavioral, and psychosocial challenges that require ongoing adaptation in care over time (Cipriani et al., 2020; Grand et al., 2011).

For many individuals, receiving a dementia diagnosis marks the beginning rather than the resolution of a complex care journey. Following diagnosis, persons living with MCI or ADRD and their care partners (together make up “dyads”) frequently experience uncertainty regarding disease progression, symptom management, treatment options, driving, legal and financial planning, available community resources, and expectations for the future (Shafir et al., 2022). As dementia progresses, informational and supportive needs evolve, requiring timely guidance that extends beyond initial diagnostic counseling (Bamford et al., 2021).

Despite growing recognition of these challenges, many dementia dyads continue to report unmet informational, emotional, and practical support needs. Families often seek answers from multiple sources—including healthcare providers, support organizations, community agencies, and the internet (Teles et al., 2022; Wasilewski et al., 2017). Yet each of these sources has important limitations. Clinical visits are typically brief and occur intermittently, leaving many questions unanswered between visits as new symptoms and caregiving challenges emerge (Bastholm-Rahmner et al., 2025; Bergqvist et al., 2026). Previous studies have also demonstrated persistent barriers to healthcare navigation, care coordination, and access to dementia-specific resources (Kern et al., 2024; Sorrentino et al., 2025). Although dementia-related information is readily available online, caregivers may have difficulty identifying information that is trustworthy, relevant, and responsive to their individual needs (Efthymiou et al., 2020). Therefore, effective support strategies are needed to assist dyads in managing the evolving challenges associated with MCI and dementia in a timely and reliable manner.

Artificial intelligence (AI) has emerged as a promising approach to augment dementia care by providing accessible, real-time, and personalized education, decision support, and resource navigation outside traditional clinical settings (Maddali et al., 2022; Yang et al., 2025). AI-based conversational tools have the potential to deliver continuous support that adapts to users’ changing needs while reducing barriers to accessing reliable information (Shi et al., 2026). However, general-purpose chatbots (e.g., ChatGPT) were not designed specifically for dementia caregiving and may provide inaccurate, overly general, insufficiently contextualized, or clinically inappropriate responses. The effectiveness and acceptability of these technologies therefore depend on user-centered development that reflects the experiences, priorities, and challenges of the individuals they are intended to support.

Developing a purpose-built AI-based chatbot for ongoing dementia care requires a comprehensive understanding of the questions, barriers, and unmet needs experienced by dementia dyads, so that the tool is clinically relevant, practical, and responsive to real-world care experiences. Although prior research has identified broad informational and caregiving needs among dementia dyads(Bamford et al., 2021; van der Roest et al., 2009), less is known about the specific questions, challenges, and support needs experienced by individuals living with MCI or early AD/ADRD and their care partners as they navigate daily life (*ADI - Dementia Statistics*, n.d.). Therefore, the purpose of this study was to explore the questions, challenges, and support needs and to use these findings to inform the development of an AI-based chatbot designed to provide ongoing support.

## Methods

### Design

A qualitative descriptive study was conducted using semi-structured interviews (Kim et al., 2017) and conventional content analysis (Hsieh & Shannon, 2005). This approach provided descriptions of experiences and perceptions of persons living with MCI or mild AD/ADRD and their care partners and informed the development of a chatbot designed to support persons living with MCI and mild AD/ADRD and their care partners by addressing common questions, challenges, needs, and support needs encountered while navigating life with MCI and dementia. Conventional content analysis (Hsieh & Shannon, 2005) was used to allow themes and subthemes to emerge directly from participant narratives. To support study translation into implementation, the Consolidated Framework for Implementation Research (CFIR) (Damschroder et al., 2022) informed development of the interview guide and provided interpretation of our findings. This study was reviewed by the institutional review board of [NAME BLINDED] and granted an exemption.

### Setting and Recruitment

Participants were recruited from academic medical centers, memory clinics, and aging organizations in the U.S., specifically in Pennsylvania, New York, Georgia, and North Carolina. Purposive sampling was used to identify persons living with MCI or mild AD/ADRD and their care partners, and dementia care experts who met study eligibility criteria and could provide rich information about the study objective. Recruitment materials included study flyers, newsletters, and referrals from collaborating organizations. Interested individuals contacted the study team and completed screening procedures. Recruitment continued until data saturation was reached. (Hennink et al., 2017)

### Study Participants

Eligible participants spoke English and were either individuals diagnosed with MCI or mild AD/ADRD and/or their informal care partners, or healthcare experts with experience providing care to persons living with cognitive impairment or dementia. The study coordinator obtained oral and written informed consent from all eligible participants. Compensation was provided for participation.

### Procedures

The study team developed the semi-structured interview guides, informed by CFIR (Damschroder et al., 2022), prior literature, and previous discussions with dementia care experts (Britt et al., 2025; Ge et al., 2025). Questions relevant to the present analysis are provided in Supplemental Files 1 and 2. The guides were designed to explore participants’ experiences living with MCI or dementia, including common questions, challenges, support needs, and opportunities for ongoing support, and were pilot-tested and refined before data collection.

The study was initially designed to focus on experiences implementing formal cognitive care plans. Early recruitment and interviews indicated that many dyads had not received a formal care plan or did not conceptualize their post-diagnosis guidance as a care plan. Eligibility and interview questions were therefore broadened to include dementia care recommendations and guidance received following diagnosis, allowing participants to describe relevant care experiences regardless of whether recommendations had been delivered as a formal cognitive care plan. The semi-structured format also allowed the interviewer to tailor probes to participants’ experiences and pursue relevant questions as they emerged.

Interviews were conducted from November 2025 to May 2026, either virtually via teleconferencing or in person according to participant preference. Dyad members were generally interviewed together; however, one care partner who was part of an enrolled dyad participated in a follow-up care-partner-only interview. The additional standalone care partner was also interviewed individually. Dementia care experts participated in individual interviews. Interviews lasted approximately 50 to 90 minutes, were audio-recorded, transcribed by a speech-to-text service, and checked for accuracy by the study interviewer/coordinator. Demographic information was also collected. Interviews continued until no additional data relevant to our study aim emerged and responses became redundant. (Hennink et al., 2017)

### Data Analysis

Interview transcripts were analyzed using conventional content analysis.(Hsieh & Shannon, 2005) Two study team members (initials blinded) independently reviewed transcript data line by line and developed preliminary codes using Dedoose software. Codes were compared, discussed, and refined through an iterative process. Similar codes were grouped into categories and subcategories reflecting participants’ experiences, questions, challenges, and support needs. The research team met regularly to discuss coding decisions, compare interpretations, and resolve any arising discrepancies, allowing discussion for framework development and refinement throughout the process. An audit trail was maintained throughout the analytic process to document coding decisions, category refinement, and consensus discussions.

Consistent with conventional content analysis, categories and themes were derived inductively from participant narratives rather than assigned to predetermined CFIR domains.(Damschroder et al., 2022) Following inductive analysis, CFIR was used as an implementation-informed interpretive lens to consider how individual and contextual factors identified in the findings may influence AI-enabled chatbot development and future implementation; findings were not coded or deductively categorized according to CFIR domains (Damschroder et al., 2022).

### Trustworthiness and Rigor

To support study methodological quality, the Consolidated Criteria for Reporting Qualitative Research checklist was used.(Tong et al., 2007) We used several strategies to enhance the trustworthiness and rigor of the findings.(Lincoln & Guba, 1985) Credibility was supported through investigator triangulation, in which study team members independently reviewed transcripts, developed preliminary codes, and met regularly to facilitate coding discussions, decisions, and category development. Any differences in interpretation were resolved through study team discussion and consensus. Credibility was further strengthened through inclusion of multiple stakeholder perspectives, including persons living with MCI or mild AD/ADRD, care partners, and dementia care experts. Dependability was enhanced through maintaining an audit trail of meeting notes, coding and category development, and analytic discussions throughout the study. Confirmability was supported through reflexive discussions among the study team to examine assumptions, interpretations, and potential biases during analysis. Transferability was facilitated by purposive sampling of dementia dyads and dementia care experts with diverse experiences across multiple geographic regions and care settings. Throughout the analytic process, the study team met regularly to conduct iterative reviews of transcripts and discuss emerging findings; this ensured that the categories accurately reflected participants’ experiences and perspectives. Further, these discussions also reflected how findings translated to informing AI Chatbot development.

## Results

### Participant Characteristics

The *dyad/care partner* sample included 11 enrolled dyads (11 persons living with MCI or mild AD/ADRD and 11 care partners) and one additional standalone care partner, for a total of 23 participants (11 persons living with MCI or mild AD/ADRD and 12 care partners) (see Table 1). Receipt of a formal cognitive care plan was not required for participation; participants could describe dementia care recommendations or guidance received following diagnosis.

**Table 1.** Sociodemographic Characteristics of Dyad/Care Partner Study Participants (n = 23)

| Characteristic | Persons living with MCI or mild AD/ADRD (n = 11) | Care Partners (n = 12) | Total sample (N = 23) |
| --- | --- | --- | --- |
| Age range | 65–97 | 35–84 | 35–97 |
| **Gender, n (%)** | — | — | — |
| Female | 4 (36.4%) | 10 (83.3%) | 14 (60.9%) |
| Male | 7 (63.6%) | 2 (16.7%) | 9 (39.1%) |
| **Race/ethnicity, n (%)** | — | — | — |
| White, Non-Hispanic | 9 (81.8%) | 10 (83.3%) | 19 (82.6%) |
| Black or African American | 1 (9.1%) | 1 (8.3%) | 2 (8.7%) |
| Hispanic or Latino | 1 (9.1%) | 1 (8.3%) | 2 (8.7%) |
| **Education, n (%)** | — | — | — |
| High school diploma/GED or less | 2 (18.2%) | 2 (16.7%) | 4 (17.4%) |
| Some college or associate degree | 1 (9.1%) | 0 (0.0%) | 1 (4.3%) |
| Bachelor's degree or higher | 8 (72.7%) | 10 (83.3%) | 18 (78.3%) |
| **Diagnosis, n (%)** | — | — | — |
| Mild cognitive impairment | 4 (36.36%) | — | — |
| Mild dementia | 7 (63.63%) | — | — |
| **Relationship to person living with MCI or mild AD/ADRD, n (%)** | — | — | — |
| Spouse/partner | — | 9 (75%) | — |
| Adult child | — | 3 (25%) | — |

Notes. GED=General Equivalency Degree

**Table 2.** Sociodemographic and Professional Characteristics of Dementia Care Experts (n = 12)

| **Characteristic** | **n (%)** |
| --- | --- |
| Age range | 30–60+ |
| **Gender, n (%)** | — |
| Female | 11 (91.7%) |
| Male | 1 (8.3%) |
| **Race/ethnicity, n (%)** | — |
| White, Non-Hispanic | 6 (50.0%) |
| Asian | 2 (16.7%) |
| Black or African American | 3 (25.0%) |
| Hispanic or Latino | 1 (8.3%) |
| **Education, n (%)** | — |
| Nursing degree | 2 (16.7%) |
| Master's degree | 4 (33.3%) |
| Doctoral degree (MD/PhD/PsyD) | 6 (50.0%) |
| **Professional role, n (%)** | — |
| Physician (geriatric, neurology, psychiatry) | 4 (33.3%) |
| Psychologist or Neuropsychologist | 2 (16.7%) |
| Social Worker | 2 (16.7%) |
| Occupational Therapist | 2 (16.7%) |
| Nurse Practitioner | 1 (8.3%) |
| Registered Nurse | 1 (8.3%) |

Persons living with MCI or mild AD/ADRD ranged in age from 65 to 97 years, and care partners ranged from 35 to 84 years. Overall, 60.9% of participants were female, 82.6% identified as White, Non-Hispanic, and 78.3% had a bachelor’s degree or higher. Among persons living with MCI or mild AD/ADRD, 36.36% were diagnosed with mild cognitive impairment and 63.63% with mild dementia. Among care partners, 75% were spouses or partners and 25% were adult children. Interview durations ranged from 45 to 120 minutes.

The *dementia care expert* sample included 12 participants representing multiple clinical disciplines. Dementia care experts ranged in age from 30 to 60+ years. Most participants were female (91.7%), and half identified as White, Non-Hispanic (50.0%). Regarding education, 50.0% held a doctoral degree, 33.3% held a master’s degree, and 16.7% held a nursing degree. Professional roles included physicians (33.3%), psychologists or neuropsychologists (16.7%), social workers (16.7%), occupational therapists (16.7%), a nurse practitioner (8.3%), and a registered nurse (8.3%).

### Findings

Three interconnected themes characterized dyads/care partners’ and dementia care experts’ experiences navigating or providing care for persons living with MCI and mild AD/ADRD: (1) questions and uncertainty emerged across multiple aspects of everyday dementia care; (2) individual, family, and healthcare system challenges complicated dementia management; and (3) ongoing support was needed as symptoms, circumstances, and care needs changed over time. Each theme comprised six subthemes reflecting the specific questions, challenges, and support needs described by participants (Figure 1). Although presented separately for clarity, the themes were interconnected and reflected the dynamic nature of navigating dementia care. Supplemental Table 1 provides the complete themes, subthemes, areas of focus, and illustrative participant quotations.

**Figure 1.**
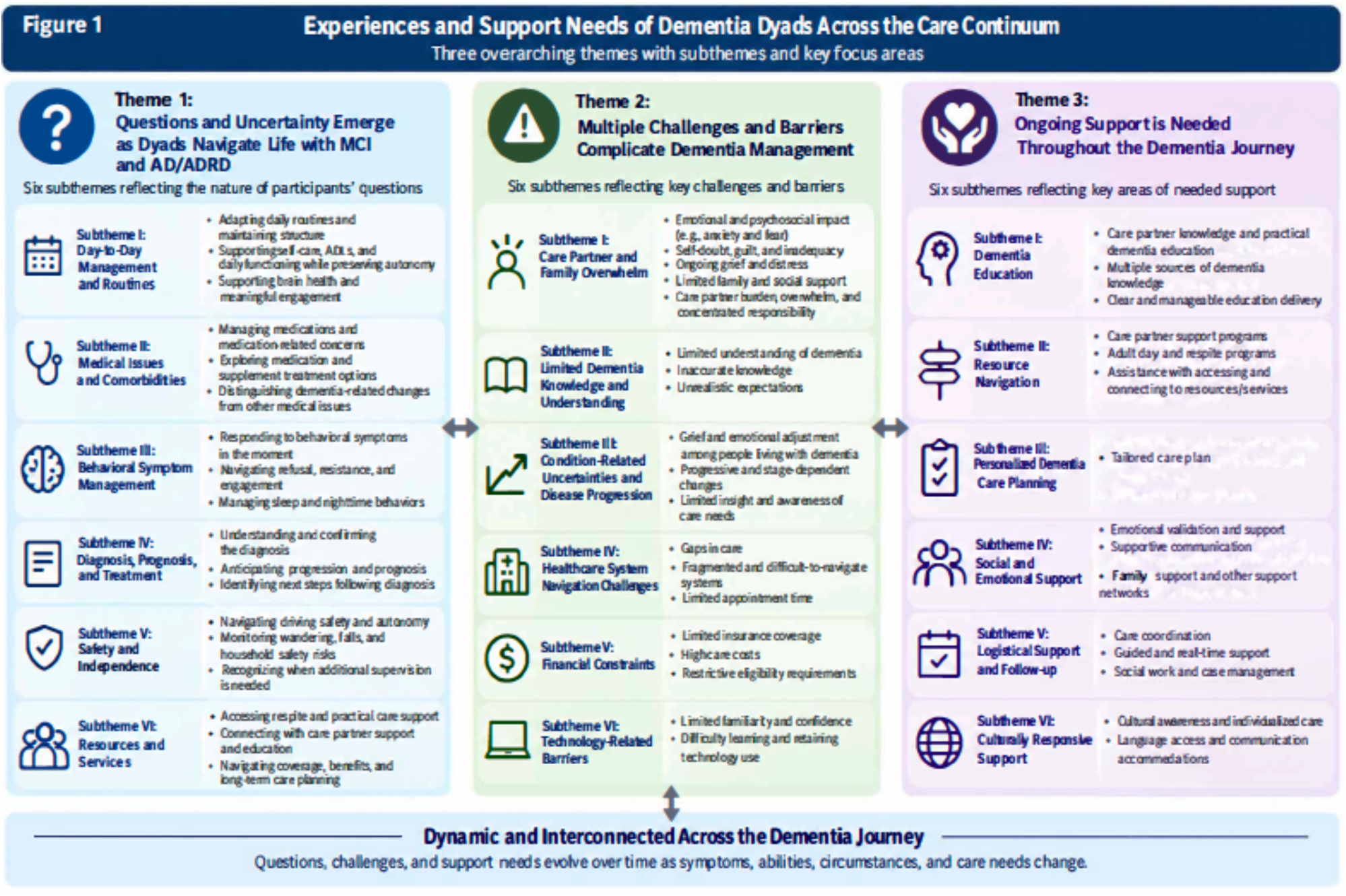
**Experiences and Support Needs of Dementia Dyads Across the Care Continuum**

#### Theme 1: Questions and Uncertainty Emerge as Dyads Navigate Life with MCI and AD/ADRD

Across interviews, dyads and dementia care experts described a broad range of questions that emerged while navigating life with MCI or AD/ADRD. Uncertainty extended beyond understanding the diagnosis and included daily routines, medical and behavioral management, safety, future changes, and access to support. These questions reflected the ongoing need to adapt as symptoms, abilities, care needs, and family circumstances changed over time. Although dyads expressed uncertainty across many areas of dementia care, they did not always know which questions to ask or what information they might need. Dementia care experts described question-asking as frequently broad or provider-led.

> *Usually, they’re not asking specific questions. It’s really up to me to just help them understand*. (Dementia care professional, Neuropsychologist)

*Day-to-Day Management and Routines*. Participants expressed uncertainty about structuring day-to-day life as abilities and support needs of the person living with MCI or mild AD/ADRD changed. Questions involved establishing routines and meaningful activities, supporting self-care and daily functioning while preserving autonomy, and incorporating nutrition, exercise, cognitive stimulation, and social engagement to promote brain health.

> *“What can I expect that they can do? What can I expect that they might need help with?” A lot of just expectation setting*. (Dementia care professional, Occupational Therapist)

*Medical Issues and Comorbidities*. Participants described uncertainty about managing medical care alongside cognitive impairment, including medication use, treatment and supplement options, distinguishing dementia-related changes from other medical conditions, and managing comorbidities. Medication management was particularly challenging when care partners were responsible for administration but were uncertain about medication purpose or benefit.

> *Why are we taking this drug? … I don’t even know what it’s doing, to be honest … it made me think, why am I giving this to him?* (Care partner 13)

*Behavioral Symptom Management*. Participants described uncertainty about responding to behavioral and emotional changes associated with cognitive impairment, including agitation, anxiety, depression, apathy, refusal or resistance to care, and changes in sleep and nighttime activity. Questions centered on practical strategies for responding in the moment while maintaining the person’s autonomy and encouraging engagement.

> *There can be defiance from what I’ve seen with regard to the individuals wanting to even do the behavioral activation activities … “I don’t want to do it.” … “What are some tactics you can think of to introduce the idea without forcing the person?”…how do you socialize the idea with them in a way that maintains their sense of agency?* (Dementia care professional, Social Worker)

*Diagnosis, Prognosis, and Treatment*. Dyads expressed uncertainty about what the diagnosis meant, how the condition might progress, and what could be done following diagnosis. Questions ranged from understanding and distinguishing dementia diagnoses and available treatments to anticipating future changes and preparing for longer-term care needs.

> *“What’s going to happen next?”… “How much longer is this person going to live, and what is life going to look like for the remaining however long that they’re here?”* (Dementia care professional, Neuropsychologist)

*Safety and Independence*. Participants described uncertainty about balancing safety with the dignity, autonomy, and independence of the person living with MCI or mild AD/ADRD. Concerns included driving, falls, wandering and household hazards, as well as determining when changes in judgment or functioning warranted greater supervision or environmental safeguards.

> *“When do you know that it’s time that I should give up driving, or when do I know that it’s time to give up doing this myself?”* (Dementia care professional, Nurse Practitioner)
>
> *If I’m not there and she falls, what’s going to happen? We don’t have, at some point, we’ll probably have to talk about some kind of alert mechanism in case she does fall when I’m not here to get some attention.* (Care partner 18)

*Resources and Services*. Participants described uncertainty about identifying and accessing services that could support both members of the dyad, including respite and in-home care, adult day programs, care partner education and support, and long-term care. Questions extended beyond what resources were available to how families could locate appropriate local services and navigate coverage, eligibility, and other requirements necessary for obtaining them.

> *“Is there anybody that you could get to come in and stay with him for a while so I could go to the grocery store?”* (Dementia care professional, Registered Nurse)

#### Theme 2: Multiple Challenges and Barriers Complicate Dementia Management

Management of MCI and AD/ADRD was shaped by challenges that reached beyond the symptoms of the condition itself. Participants described emotional strain within families, gaps in dementia knowledge, uncertainty as the condition progressed, difficulty accessing and coordinating care, financial limitations, and barriers to using technology-based supports. These challenges often interacted, creating additional demands for dyads as they attempted to make decisions, respond to changing needs, and obtain appropriate support.

#### Care partners and Family Overwhelm

Care partners described substantial emotional and practical strain as caregiving responsibilities accumulated and the needs of the person living with MCI or mild AD/ADRD changed. Anxiety, fear, self-doubt, guilt, and grief occurred alongside limited family and social support, leaving some care partners carrying substantial responsibility with little assistance. Caregiving demands often competed with employment, household responsibilities, personal obligations, and care partners’ own health and emotional needs.

> *I feel like I have to do everything like…I got to entertain, make sure the healthcare, all the needs are met and also work and deal with my own grief*. (Care partner 5)

Together, emotional strain, limited support, and concentrated responsibility contributed to a sense of overwhelm and made caregiving more difficult to sustain.

> *I think the main thing is caregivers are just so underwater. There’s just so much that they have to do and they’re so stressed. And so, everything that then it’s even such a big part of this too, is not having time for themselves. They don’t have enough support as we talked about. They don’t have enough support financially, legally, emotionally, and socially*. (Dementia Care Professional, Social Worker)

#### Limited Dementia Knowledge and Understanding

Gaps in foundational and accurate dementia knowledge made it difficult for care partners to interpret cognitive, behavioral, and functional changes, adjust expectations, and apply care recommendations. Participants described limited understanding of dementia beyond memory loss, including instances in which dementia-related changes were interpreted as intentional behavior or personality characteristics and expectations that functioning might improve despite disease progression.

> *I think the challenges come from me because I’m not understanding the decline or not accepting it maybe*. (Care partner 13)
>
> *I think of the patients where their family member is convinced that they are just stubborn and they can’t wrap their head around the fact that they actually have a brain condition.* (Dementia Care Professional, Geriatrician)

#### Condition-Related Uncertainties and Disease Progression

Participants described challenges adapting to the progressive and changing nature of dementia, including emotional adjustment to declining abilities, stage-dependent changes in care needs, and differences in awareness of cognitive impairment within the dyad. Persons living with MCI or mild AD/ADRD described frustration, loneliness, and grief as changes affected their independence and sense of self.

> *I’m told it’s not going to change… I have abilities, but I just am not doing them as well as I did…it’s frustration and its definitely loneliness. I was better at a multitude of things. I can’t. (Person living with MCI or mild AD/ADRD 13)*
>
> *One of the hardest things is the patient has no idea that they have cognitive impairment when the family member totally understands what needs to happen…the disconnect of patients who have dementia and have support but are so unwilling to receive that support… there’s just this massive gap between what their care should look like and what they will let happen to them.* (Dementia Care Professional, Geriatrician)

#### Healthcare System and Navigation Challenges

Participants described difficulty obtaining timely, coordinated, and responsive dementia care due to gaps in follow-up, fragmented referral pathways, long waits for services, and limited time during clinical encounters. Families sometimes felt that concerns were not adequately recognized or addressed and were left to navigate multiple providers and services with limited coordination.

> *Sometimes we’ll make treatment plans, but I don’t think a lot of it is realistic for them to do…I see the referrals were marked neuropsychologists. I mean, that’s like a one-year wait list. So, what’s that really going to do? Vision screening, hearing screening…There’s just a lot of communication lost because there’s no centralized language.* (Dementia Care Professional, Geriatric Psychiatrist).

#### Financial Constraints

Financial barriers limited families’ access to dementia-related care and support through gaps in insurance coverage, high out-of-pocket costs, and restrictive eligibility requirements. Services such as medications, in-home assistance, respite, therapies, and residential care were sometimes unavailable or unaffordable, while some families did not qualify for assistance despite being unable to pay privately.

> *I’ve had patients who have not been able to be on medications just because their insurance changed and they couldn’t find a provider who took their insurance and they were off medications for a year and a half.* (Dementia care professional, Neuropsychologist)

#### Technology-Related Barriers

Technology created additional barriers to accessing dementia-related information, services, and communication. Some participants had limited familiarity or confidence with digital tools, while cognitive changes could make learning new technologies, remember steps, or continue to use previously familiar technologies more difficult.

> *I had to learn a whole new software and it was hard. It was really, really hard…I used to do all that kind of stuff and suddenly it just doesn’t.* (Person living with MCI or mild AD/ADRD 14)

#### Theme 3: Ongoing Support Is Needed Throughout the Dementia Journey

Participants identified a range of supports needed to address evolving questions and challenges associated with MCI and dementia. Desired support extended beyond clinical treatment and included education, resource navigation, individualized planning, emotional and social support, care coordination, and culturally responsive services. Across these areas, participants emphasized the importance of continued access to guidance as symptoms, circumstances, and care needs changed over time.

#### Dementia Education

Participants emphasized the need for ongoing education to help care partners understand dementia symptoms and progression, anticipate changing needs, and respond to behavioral, communication, and safety concerns. Care partners described obtaining information through healthcare professionals, formal education, support groups, other care partners, and community resources. Importantly, participants emphasized that the usefulness of education depended not only on its content but also on how it was delivered; information needed to be clear, understandable, prioritized, and provided in manageable amounts.

> *It just seems like so much information that as long as it came in smaller chunks, it would be less challenging.* (Dementia Care Professional, Geriatrician)

#### Resource Navigation

Participants emphasized that community and care partner resources were important extensions of dementia care, including support groups, care partner programs, adult day and respite services, and other community-based supports. These resources provided practical guidance, social connection, meaningful engagement, and respite; however, availability alone was insufficient. Dyads also needed help identifying which services fit their needs and connecting with appropriate supports.

> *I think attending the memory cafes have helped. They keep my mind going. It’s always something new and you’re still learning. I think that being able to focus and learn are helpful…we do artwork and I love doing artwork, so that helps too. That would be the most beneficial.* (Person living with MCI or mild AD/ADRD 20)

#### Personalized Care Planning

Participants emphasized that dementia care planning should be individualized and flexible rather than based on a fixed set of recommendations. Plans needed to account for the person’s abilities, routines, preferences, goals, disease stage, and changing circumstances while also addressing everyday functioning and anticipating future care needs.

> *It’s hard to give a recipe to someone and be like, do this and this and this, and then that will work. It really is dependent on the person.* (Dementia Care Professional, Geriatrician)

#### Social and Emotional Support

Participants emphasized the importance of social and emotional support for both persons living with MCI or mild AD/ADRD and their care partners. Support included patient, reassuring, and validating communication with the dyad; practical and emotional assistance from family, friends, and broader social networks; and opportunities to express difficult emotions and feel understood without judgment. For persons living with MCI or mild AD/ADRD, meaningful support also involved a sense of mutual recognition and human connection.

> *I want somebody who looks at me that I could see that maybe they have a hardship themselves somewhere. And then you know what? It’s like eye to eye.* (Person living with MCI or mild AD/ADRD 13)

#### Logistical Support and Follow-up

Participants highlighted the need for practical support that extended beyond individual clinical encounters and helped families translate care recommendations into action. This included coordination across providers and services, timely guidance when questions or unexpected situations arose, and social work or care management assistance with accessing resources, navigating healthcare and financial systems, and planning for future needs. Integrated care reduced the burden of managing disconnected services.

> *I think there’s just a level of relaxation now with this new clinic…Versus trying to catch all the different doctors…It’s all under one roof.* (Care partner 13)

#### Culturally Responsive Support

Dementia care experts emphasized that support be responsive to individuals’ cultural identities, beliefs, values, and language needs rather than assuming a uniform approach across families. This included considering cultural and religious beliefs when planning care and ensuring that dementia assessment, education, and communication were provided in a language dyads and families could understand, including through bilingual providers or interpreters when needed.

> *Like a religion is addressed as well because there are religions that don’t allow for blood transfusions. There are religions that don’t allow for other things. So, we have to be aware of that whenever we’re planning their care.* (Dementia care professional, Registered Nurse)

#### Translation of Findings to AI-Enabled Support Design

To inform the development of an AI-enabled chatbot for ongoing dementia care, findings were considered in relation to opportunities for providing ongoing support outside traditional clinical encounters (Table 3). Persistent and evolving questions highlighted the need for a comprehensive, clinically organized, expert-vetted knowledge base, so that the chatbot can provide accessible, trustworthy guidance when concerns arise. The findings in Theme 1 directly informed the development of this knowledge base. The challenges described by participants underscored the importance of presenting information in clear, manageable, and actionable ways. Identified support needs further indicated opportunities for personalized education, resource navigation, and guidance responsive to changing circumstances. These findings informed design priorities for the chatbot while also identifying essential needs, such as human connection, clinical care, and community-based services, that AI-enabled support should complement, not replace.

**Table 3.** Translation of Qualitative Findings into Design Priorities for AI-Enabled Dementia Support.

| Empirically Derived Finding | What the Finding Indicates | AI-Enabled Support Design Priority |
| --- | --- | --- |
| Questions and Uncertainty Emerge as Dyads Navigate Life with MCI and Dementia | Dyads encounter evolving questions about daily management, medical concerns, behavioral symptoms, diagnosis/prognosis, safety, and available resources; they may not always know what questions to ask. | **Accessible, on-demand guidance**, drawn from a comprehensive, clinically organized, expert-vetted knowledge base, providing trustworthy information and practical responses to questions as they arise |
| Multiple Challenges and Barriers Complicate Dementia Management | Overwhelm, limited dementia knowledge, disease-related uncertainty, healthcare navigation difficulties, financial constraints, and technology barriers can complicate families' ability to identify and act on appropriate next steps. | **Clear, manageable, and actionable support** that simplifies information, assists with decision-making and navigation, and minimizes additional cognitive or technological burden. |
| Ongoing Support is Needed Throughout the Dementia Journey | Dyads need education, resource navigation, personalized care planning, social/emotional support, logistical follow-up, and culturally responsive support as needs and circumstances change. | **Personalized and responsive ongoing support** that connects users with relevant education and resources and provides guidance between healthcare encounters. |

*Note. Themes and subthemes were derived inductively through conventional content analysis. Design priorities reflect the translation of participants' identified questions, challenges, and support needs into considerations for development of the AI-enabled chatbot. AI-enabled support is intended to complement clinical care, human support, and community-based services, not replace them.*

## Discussion

In this qualitative descriptive study, we explored the questions, challenges, and support needs experienced by persons living with MCI or mild AD/ADRD, their care partners, and dementia care experts. Findings demonstrate that post-diagnostic dementia management is characterized by evolving uncertainty, multilevel barriers to managing care, and support needs that extend beyond episodic clinical encounters. Importantly, these findings were interconnected: questions emerged as dyads encountered new situations, individual and system-level barriers complicated their ability to respond, and participants identified needs for ongoing, personalized, and practical support. Together, the findings suggest that post-diagnostic dementia support should be conceptualized as a responsive process that helps dyads navigate changing needs in everyday life rather than as a discrete period of education following diagnosis

### Questions Persist Throughout the Dementia Journey

A central finding of this study was that questions and uncertainty persisted beyond diagnosis and extended across both clinical and everyday aspects of living with cognitive impairment. Dyads sought information about diagnosis, prognosis, and treatment while simultaneously navigating practical concerns related to daily routines, behavioral symptoms, medical management, safety, independence, and available resources. Rather than resolving uncertainty, diagnosis appeared to mark the beginning of an ongoing process of information seeking as individuals and families encountered new situations and attempted to understand what cognitive impairment meant for their daily lives. This finding is consistent with prior research demonstrating that the information needs of persons living with MCI or mild AD/ADRD and their care partners are broad and evolve as dementia progresses and caregiving responsibilities change (Soong et al., 2020). Our findings extend this work by illustrating the range of questions that may arise even during relatively early stages of cognitive impairment and by demonstrating how disease-related questions coexist with highly practical concerns about managing everyday life.

Importantly, participants also described that dyads did not always know what information they needed or which questions to ask. This finding suggests that post-diagnostic support cannot rely solely on individuals and families to recognize emerging needs and initiate information seeking. The relevance of particular information may also change over time: information about diagnosis and prognosis may be salient following diagnosis, whereas guidance about behavioral symptoms, safety, or care services may become more meaningful when those situations are encountered in daily life. Prior research similarly suggests that care partners’ preferences for the type and timing of dementia information vary across the dementia care trajectory (Berghout et al., 2024). Providing large amounts of information at diagnosis may therefore be insufficient if families do not yet recognize its relevance or are unable to anticipate the situations in which it will be needed.

These findings suggest that post-diagnostic information should be available over time and responsive to new questions and changing needs, rather than provided primarily at diagnosis. Dyads may benefit from opportunities to revisit information and obtain practical guidance as new questions arise, abilities change, and care demands evolve. Individualized post-diagnostic follow-up and access to guidance beyond the initial diagnostic encounter may be particularly important for helping families translate clinical information into everyday dementia management (Caprioli et al., 2023; Ng et al., 2016). This approach shifts the emphasis from providing all potentially relevant information at one time point to making trustworthy and relevant guidance accessible over time as it becomes meaningful to the dyad.

### Individual and System-Level Barriers Complicate Dementia Management

Our findings demonstrate that difficulties managing MCI and dementia cannot be solely understood as simply dementia knowledge deficits. Participants described interconnected challenges operating across the individual, family, healthcare-system, and resource contexts, including care partner overwhelm, limited dementia knowledge, uncertainty associated with disease progression, fragmented healthcare systems, financial constraints, and technology-related barriers. These challenges frequently intersected with the questions dyads were attempting to resolve. For example, recognizing that additional assistance was needed did not necessarily mean that a family could identify an appropriate service, navigate referral processes, qualify for assistance, afford available care, or access information through digital platforms. Thus, what initially appears to be an information need may also reflect practical and structural barriers that further constrain families’ ability to act on available information.

These findings are consistent with literature demonstrating that dementia caregiving requires substantial healthcare-management work in addition to direct caregiving, including coordinating appointments and services, communicating with healthcare professionals, navigating insurance and benefits, and participating in treatment and care decisions (Mueller et al., 2022; Zuber et al., 2026). Our findings further illustrate how these demands can accumulate alongside emotional strain, employment and household responsibilities, and care partners’ own health needs. Limited family and social support may further compound these responsibilities within one care partner. Consequently, interventions that focus primarily on increasing dementia knowledge alone may have limited impact as care partners simultaneously lack the time, resources, coordination, or practical assistance necessary to implement recommendations.

Viewed through an implementation lens, the findings also illustrate influences operating across individual and contextual levels. Limited dementia knowledge and care partner overwhelm may shape families’ ability to recognize changing needs and determine appropriate responses, while service availability, financial constraints, fragmented referral pathways, and limited follow-up may shape whether needed support can actually be accessed. CFIR provided a useful lens for considering these multilevel influences without determining the inductively derived themes (Damschroder et al., 2022). These findings therefore suggest that barriers encountered by dementia dyads arise not only from individual knowledge or caregiving capacity but also from the broader systems through which dementia care and supportive services are accessed.

This distinction has important implications for intervention development. Information alone is unlikely to address support needs when families also face fragmented services, financial constraints, and difficulty determining how to navigate available care. Dementia support interventions may therefore need to combine education with practical strategies that help dyads identify appropriate resources, navigate services, determine next steps, and connect with human or professional assistance when needed (Kallmyer et al., 2023; Kokorelias et al., 2023). Such approaches move beyond simply making information available toward helping families use that information within the realities of everyday dementia care context.

### Ongoing Support Needs Extend Beyond Clinical Encounters

Participants’ descriptions of needed support reinforced that post-diagnostic dementia care involved more than providing additional information. Dyads identified needs for dementia education, resource navigation, personalized care planning, logistical assistance and follow-up, culturally responsive care, and social and emotional support. Collectively, these findings suggest that ongoing support must help dyads not only understand dementia but also apply information, identify and access appropriate resources, plan for changing needs, and respond to challenges as they emerge. This multidimensional view is consistent with previous research demonstrating that the needs of persons living with MCI or mild AD/ADRD and their care partners evolve throughout the disease trajectory and require individualized and ongoing support (Bressan et al., 2020; Caprioli et al., 2023).

Across these support domains, participants also highlighted characteristics that may make support more usable in everyday life. Participants noted that education needs to be understandable, prioritized, and delivered in manageable amounts rather than overwhelming families with information. Care planning needed to reflect individual abilities, routines, preferences, goals, and disease stage rather than relying on uniform recommendations. Similarly, identifying resources was not sufficient when families did not know which services were relevant or how to connect with them. These findings highlight an important distinction between support availability and support usability: services and information may exist without being readily actionable for families. Dementia care-navigation models similarly emphasize tailored support that connects families with appropriate medical, social, financial, and community resources while integrating education and care coordination (Kallmyer et al., 2023).

The need for continuity was also displayed in participants’ responses about care coordination, real-time guidance, social work and case management, and assistance when circumstances change. Many of the questions described in this study arose unpredictably in everyday life rather than during scheduled healthcare encounters. Although clinical professionals remain central to diagnosis, treatment, and care planning, individual appointments may be insufficient to address needs that emerge between visits. Complementary mechanisms for providing tailored education, care navigation, and follow-up may therefore help bridge gaps between clinical encounters and support families as they attempt to implement recommendations at home.

Finally, participants’ emphasis on social and emotional support underscores that effective dementia support cannot be reduced to information, navigation, or care coordination. Dyads expressed needs related to reassurance, validation, supportive communication, social connection, and assistance from broader support networks. These relational dimensions are particularly important when considering technology-enabled approaches to dementia support: some needs may be facilitated by improved access to information or navigation, whereas others fundamentally depend on human relationships, professional expertise, and community-based services. Altogether, the findings support a person- and family-centered approach in which ongoing support complements clinical encounters and adapts to the informational, practical, and emotional needs of dyads as those needs change over time (Caprioli et al., 2023; Kallmyer et al., 2023).

### Implications for AI-Enabled Dementia Support

Findings from this study inform user-centered guidance for the development of AI-enabled tools intended to support dementia dyads between clinical encounters. The potential role for conversational tools may provide much-needed on-demand access to trustworthy dementia information and practical guidance as questions evolve and concerns arise. Participants’ emphasis on clear and manageable education further suggests that AI-enabled support should prioritize relevant information and present it in a concise, digestible format rather than overwhelming users with broad or generic recommendations. Similarly, findings related to resource navigation and personalized care planning suggest opportunities for AI-enabled tools to help dyads identify relevant resources, organize next steps, and access guidance responsive to their individual circumstances and changing needs.

At the same time, our findings identify important boundaries for AI-enabled dementia support tools. Technology-related barriers, including familiarity with digital tools and cognitive changes affecting technology use, may limit accessibility for some users and underscore the importance of simple, cognitively accessible design. Moreover, participants reported needs for human interaction, emotional validation, clinical judgement, care coordination, and community-based services that cannot be replaced by a chatbot. AI-enabled support may therefore be most appropriately positioned as a complementary resource that extends access to education, practical guidance, and resource navigation while facilitating connection to health professionals and other human supports when needed. These considerations are informing development of the AI-based chatbot in the broader study and will guide subsequent usability and acceptability testing.

### Strengths and Limitations

This study has several strengths. Inclusion of persons living with MCI or mild AD/ADRD, care partners, and multidisciplinary dementia care experts provided multiple perspectives on questions, challenges, and support needs encountered in dementia care. Including persons living with MCI or mild AD/ADRD is essential for ensuring that their experiences and priorities are captured and represented accurately, alongside care partner and professional perspectives. Participants were recruited across multiple U.S. geographic regions and care settings, and dementia care experts represented diverse clinical disciplines. The qualitative descriptive design and conventional content analysis also allowed findings to be derived inductively from participants’ accounts, while investigator triangulation, iterative team-based analysis, and maintenance of an audit trail supported analytic rigor.

Several limitations should be considered. Our study focused on individuals with MCI or mild dementia; therefore, findings may not reflect questions and support needs that become more prominent in moderate or advanced dementia. The dyad sample was predominantly White, non-Hispanic (82.6%) and highly educated, with 78.3% reporting a bachelor’s degree or higher, which may limit transferability to populations with greater racial, ethnic, educational, linguistic, and socioeconomic diversity. Participants were English-speaking, further limiting representation of individuals who may experience language-related barriers to dementia care. Because recruitment materials described a study involving development of a technology-based support tool, individuals willing to participate may also have differed from the broader population in their interest in or comfort with technology. Finally, these qualitative findings identify perceived needs and potential design considerations but do not establish the effectiveness of any specific support strategy or AI-enabled intervention.

### Future Directions

Our findings are informing the next phases of AI-enabled chatbot development, including refinement of its educational content, personalization, resource-navigation functions, and approaches for providing clear and manageable guidance. Subsequent work should evaluate the usability and acceptability of the chatbot with persons living with MCI or dementia and their care partners, with particular attention to cognitive accessibility and technology-related barriers identified in this study.

Future directions should examine whether the questions and support needs identified here are represented among more diverse populations and later stages of dementia. Studies including greater racial, ethnic, linguistic, socioeconomic, educational, geographic (including rural communities), and technological diversity will be important for understanding how support preferences and barriers vary across communities. Additional research should examine how support needs evolve longitudinally as dementia progresses to determine which needs are appropriately addressed through digital tools, healthcare, community-based, and interpersonal supports. Ultimately, evaluation of AI-enabled dementia support should consider not only usability and engagement but also whether these tools improve access to trustworthy information and resources without increasing burden or widening existing disparities in dementia care.

## Conclusions

Dementia dyads experience evolving questions and interconnected informational, practical, emotional, and system-level challenges as they navigate life following diagnosis. Findings demonstrate that post-diagnostic support requires more than information provision alone; dyads need ongoing, personalized, and actionable support that helps them understand changing needs, navigate available resources and services, plan for future care, and obtain assistance as challenges emerge in everyday life. Support must also remain responsive to the social, emotional, and relational dimensions of living with and caring for someone with cognitive impairment. AI-enabled tools may offer one complementary approach for extending access to education, practical guidance, and resource navigation between clinical encounters, but should be designed to augment, not replace clinical care, human connection, and community-based support.

## Data Availability

The anonymous transcripts of the interviews can be viewed upon request. Please send requests
to the following e-mail address

## Acknowledgements

The authors thank all participants and research staff for their contribution to data collection.

## Consent Statement

All human subjects provided informed consent for this study.

## Data Availability

The anonymous transcripts of the interviews can be viewed upon request. Please send requests to the following e-mail address

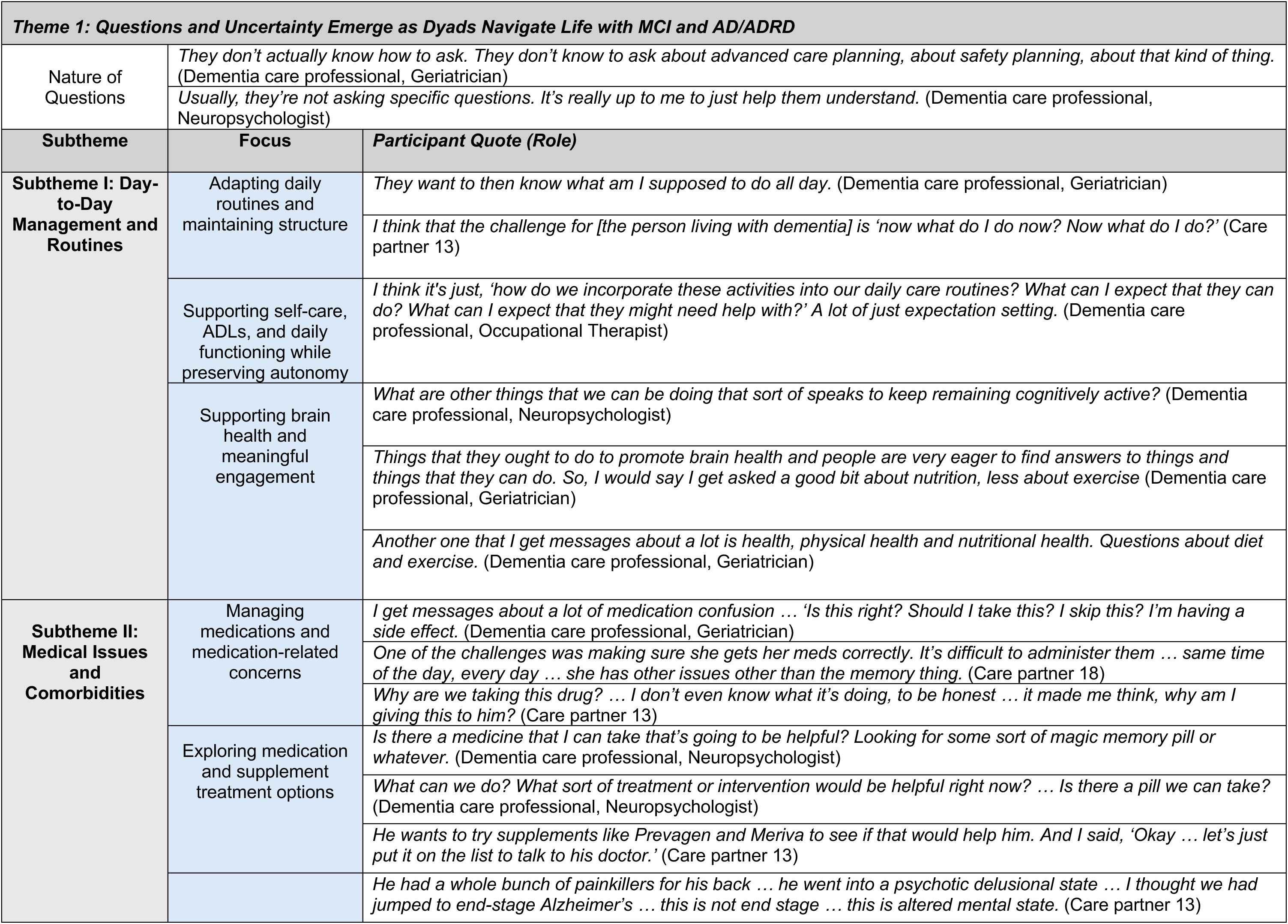
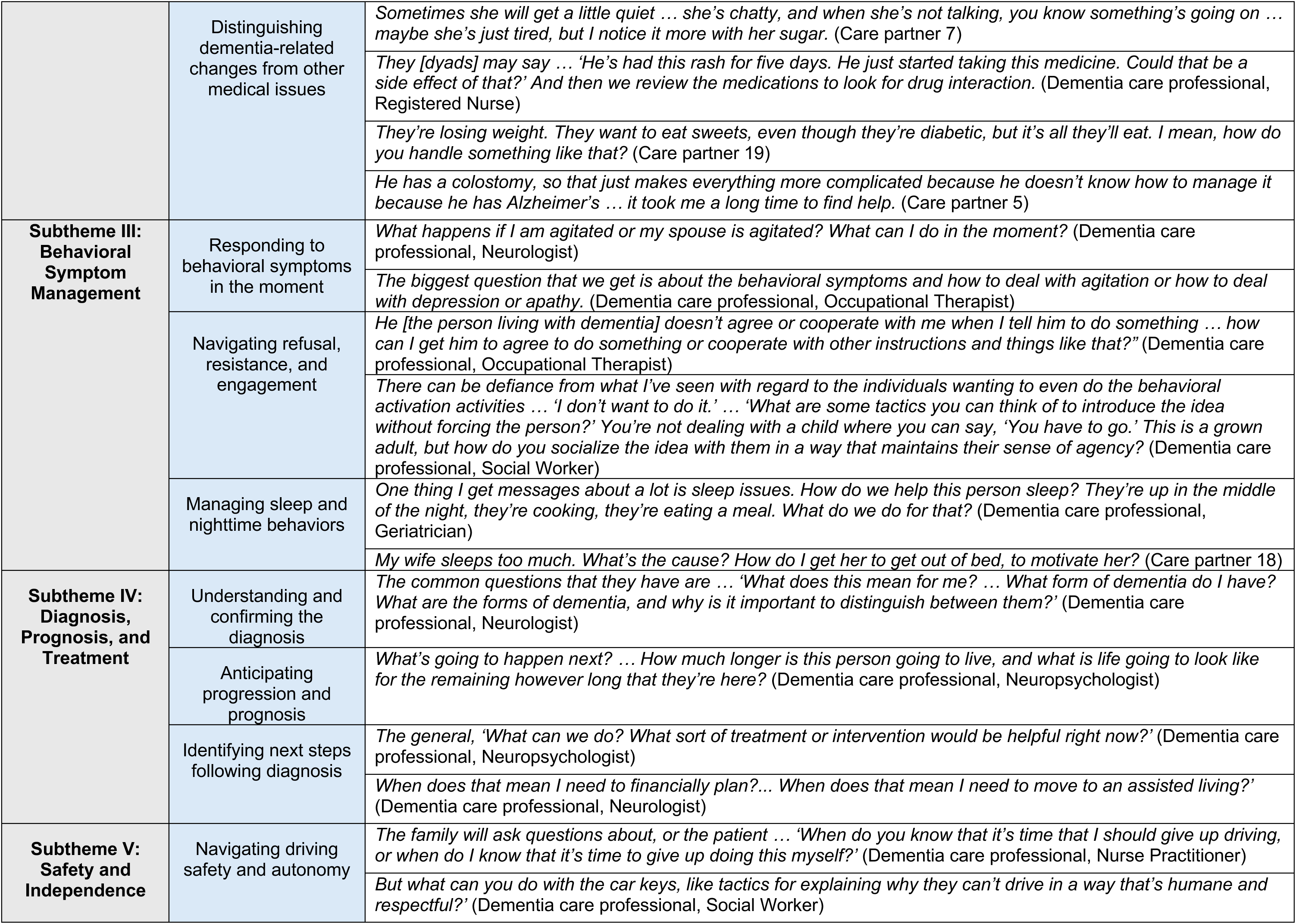
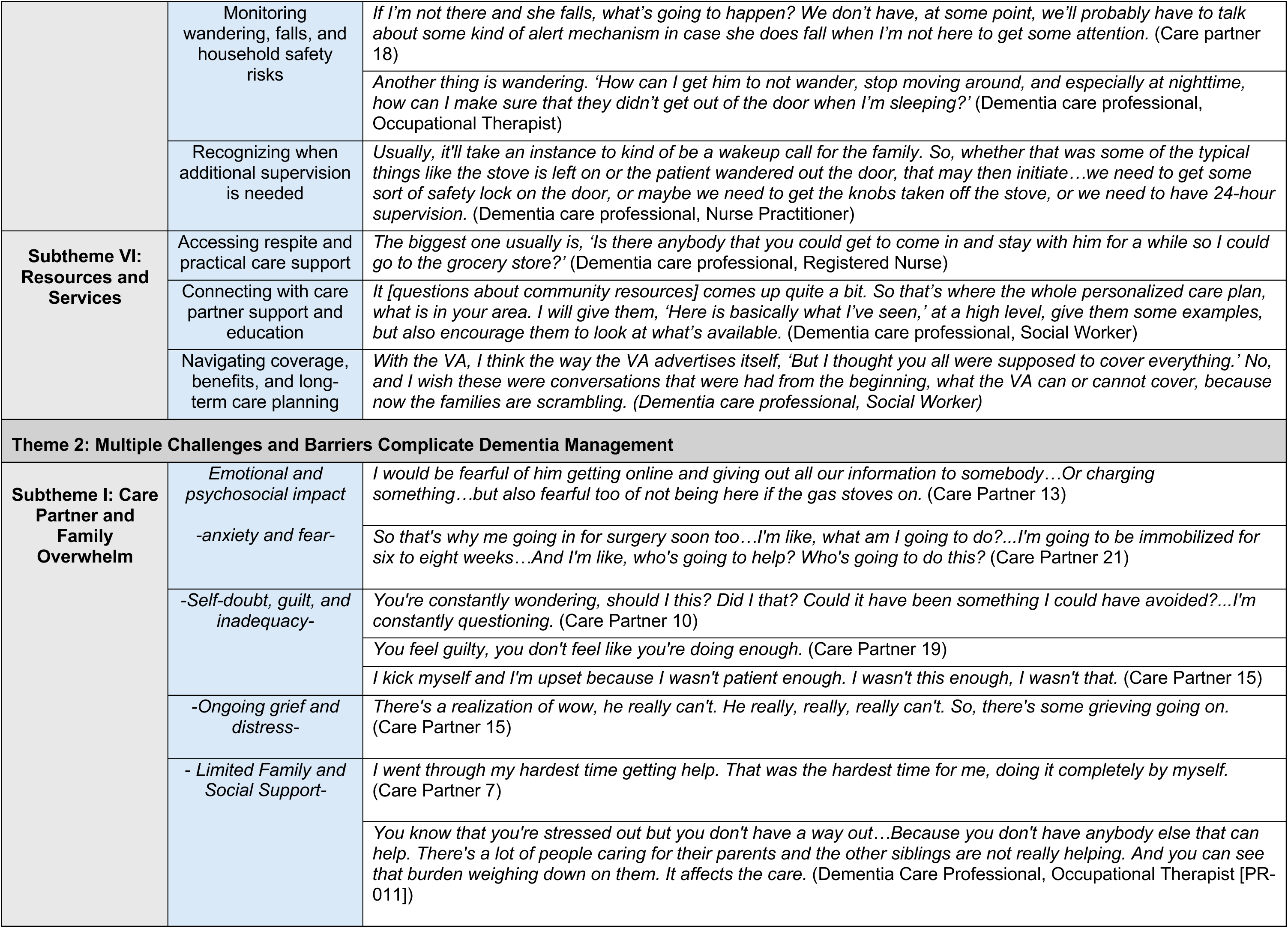
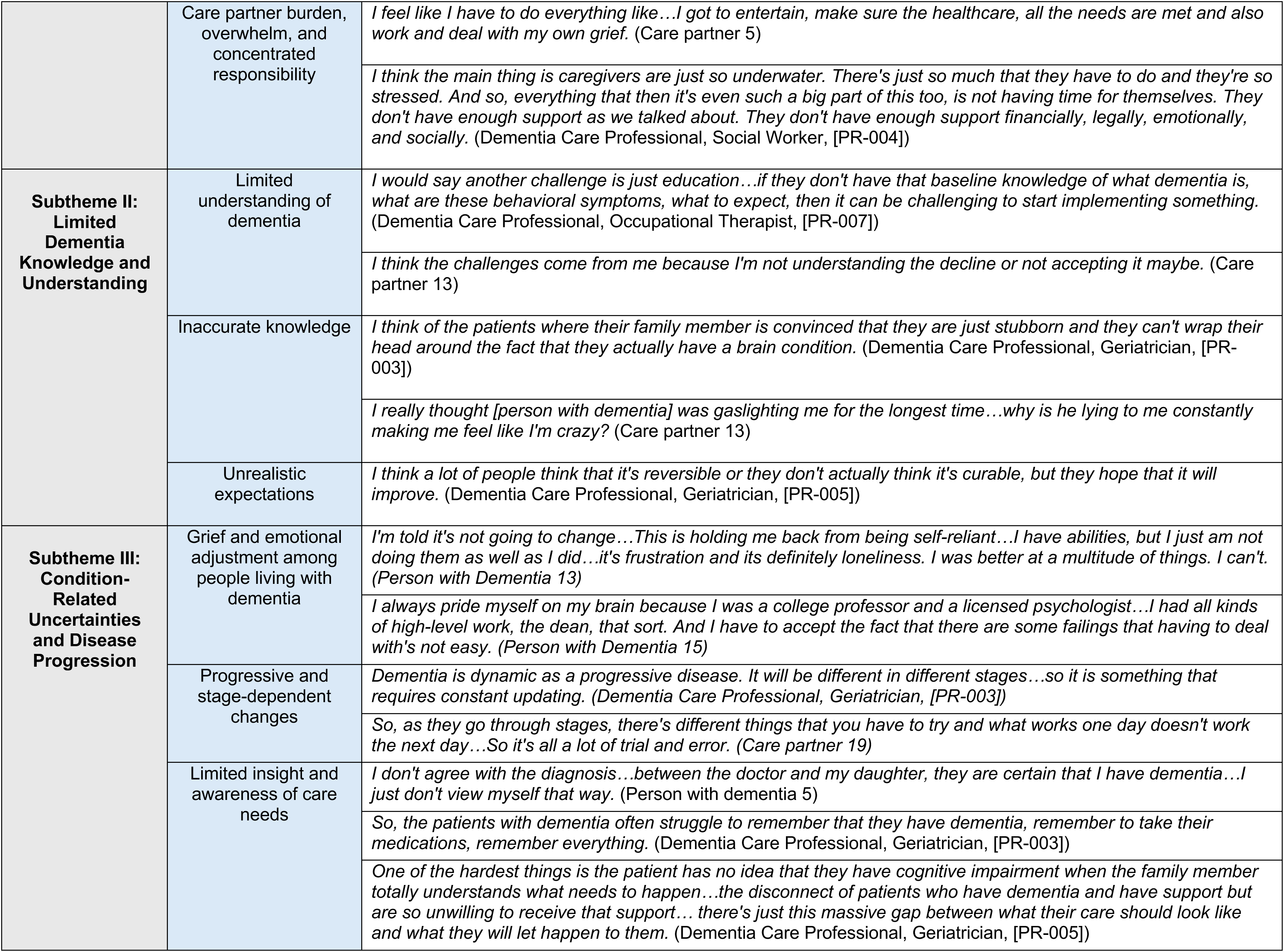
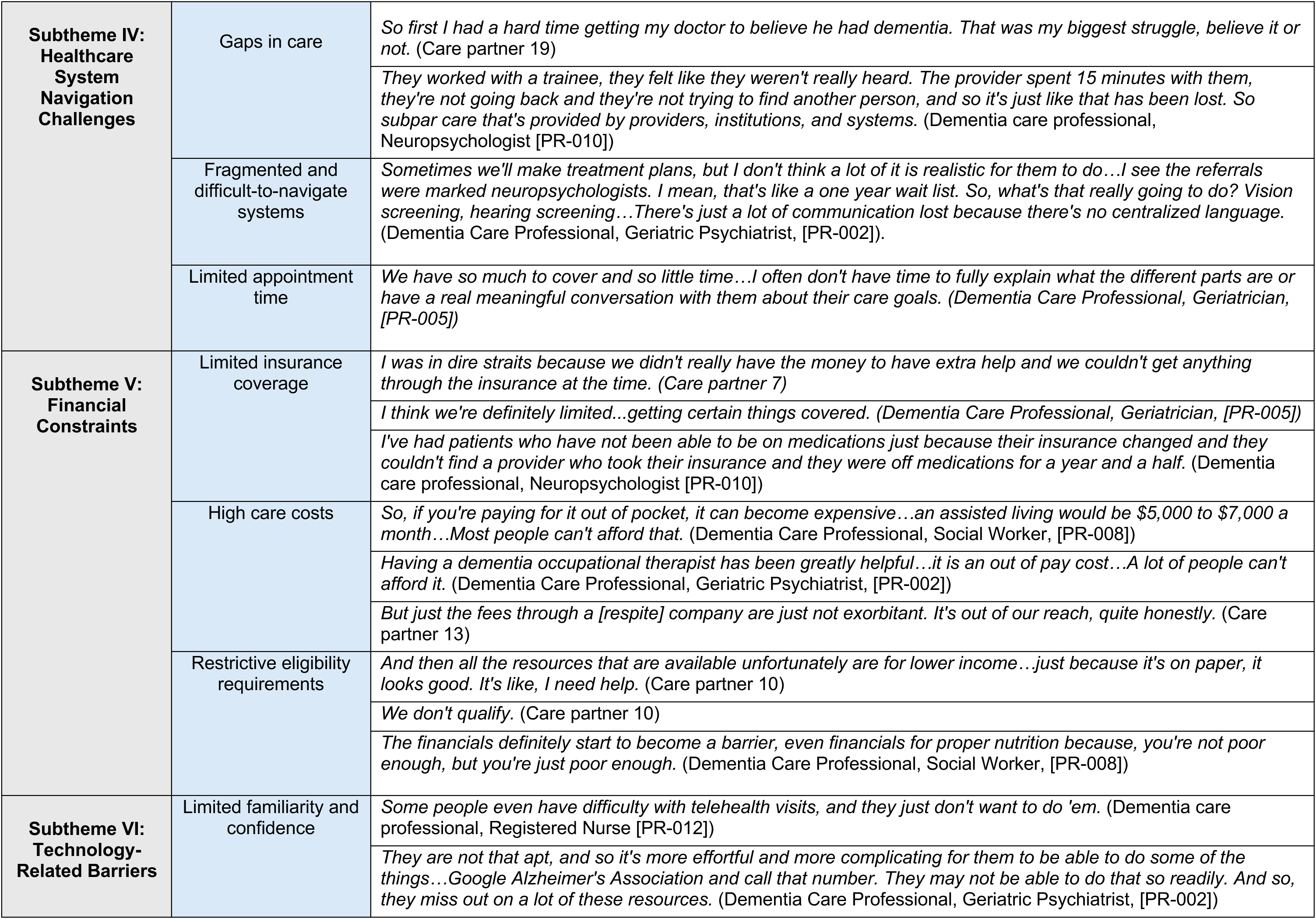
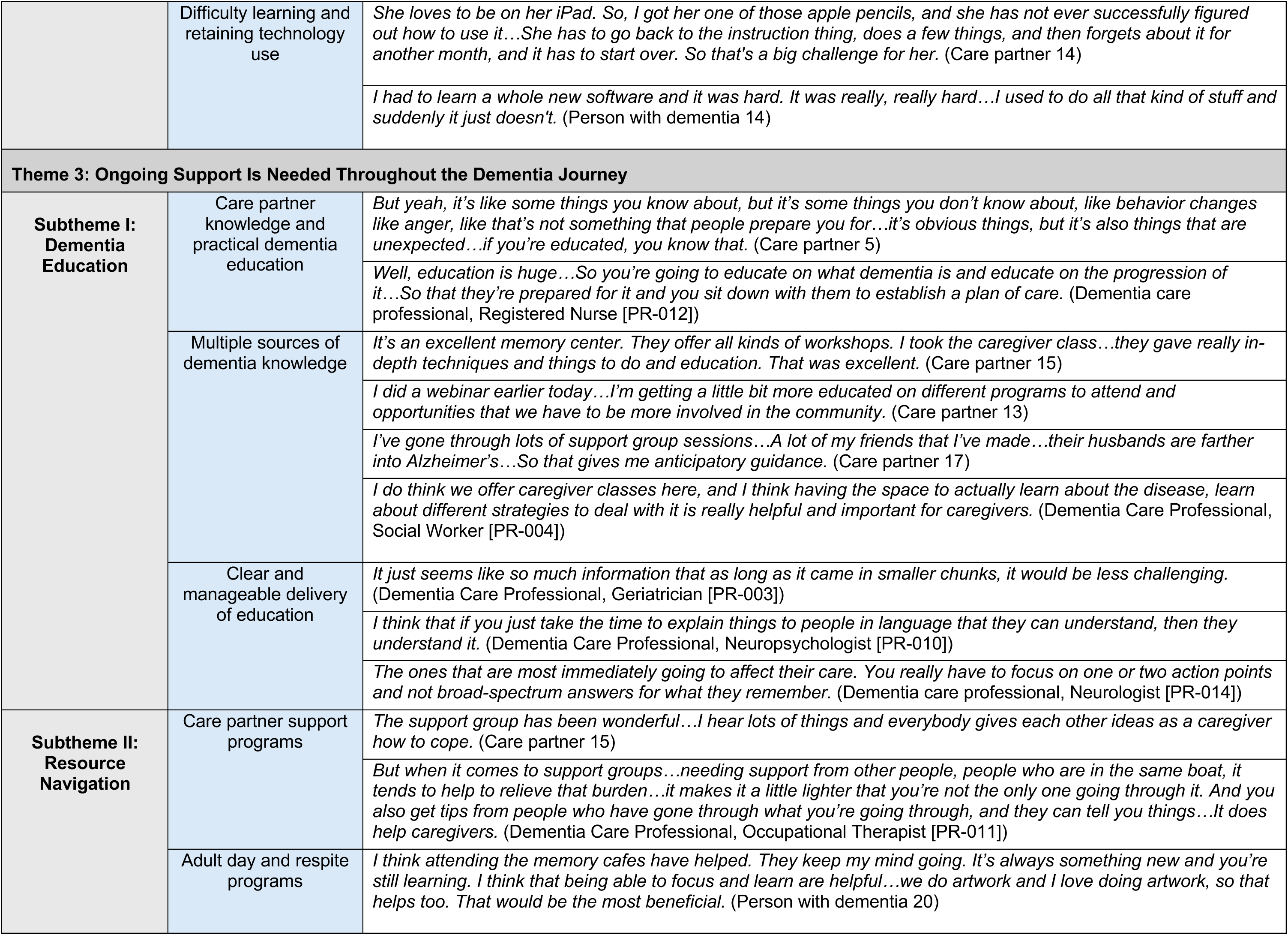
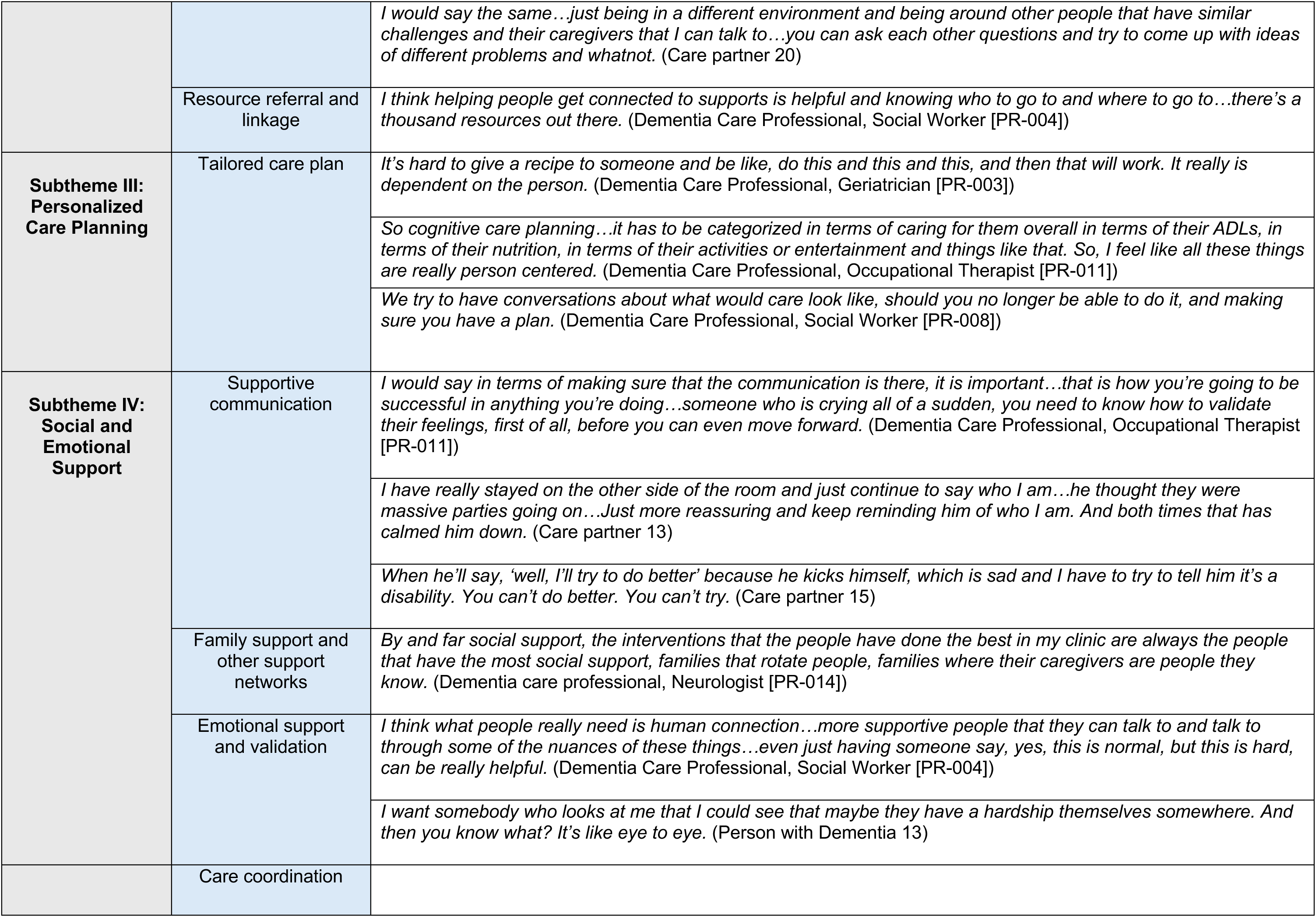
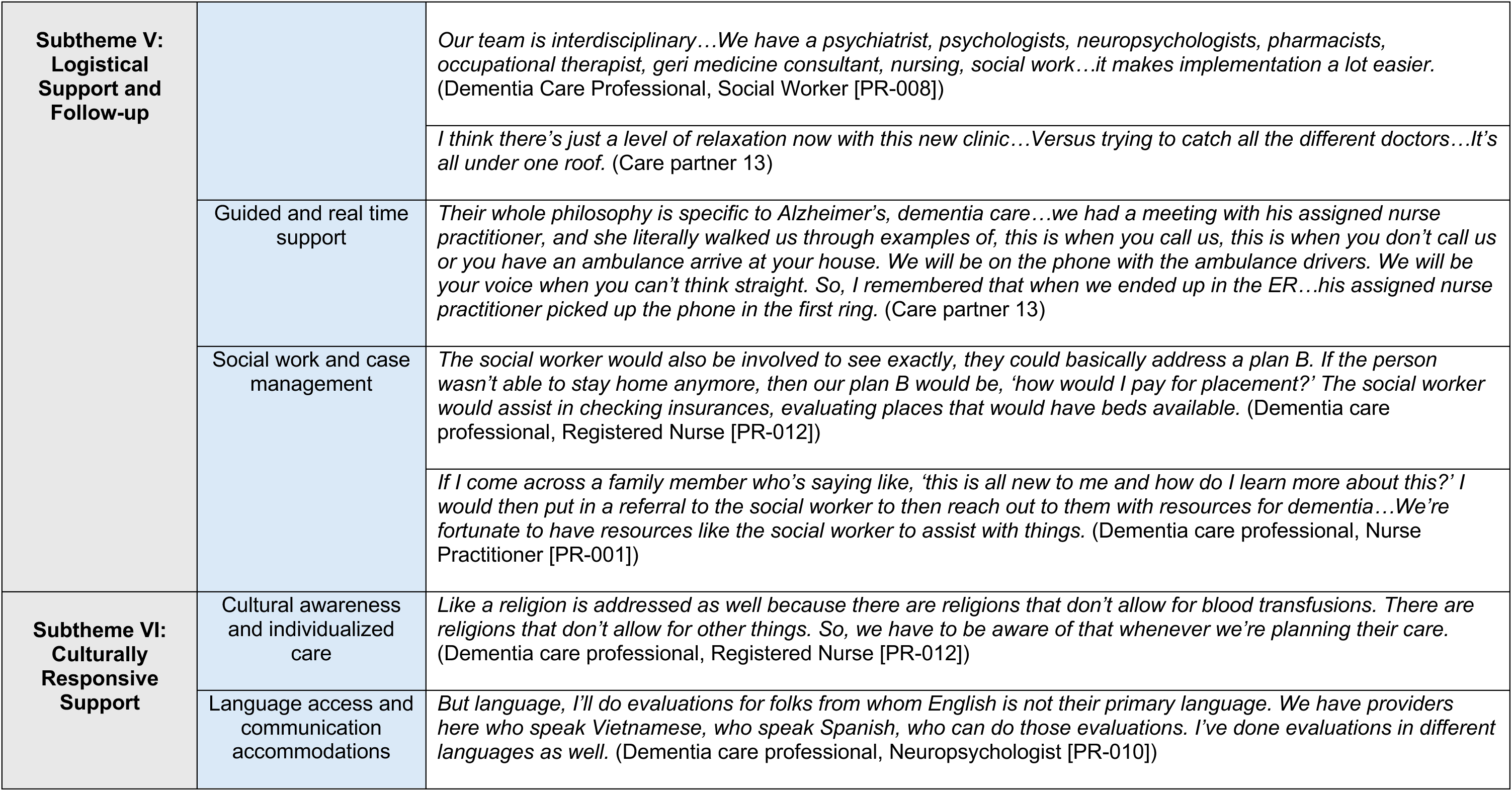
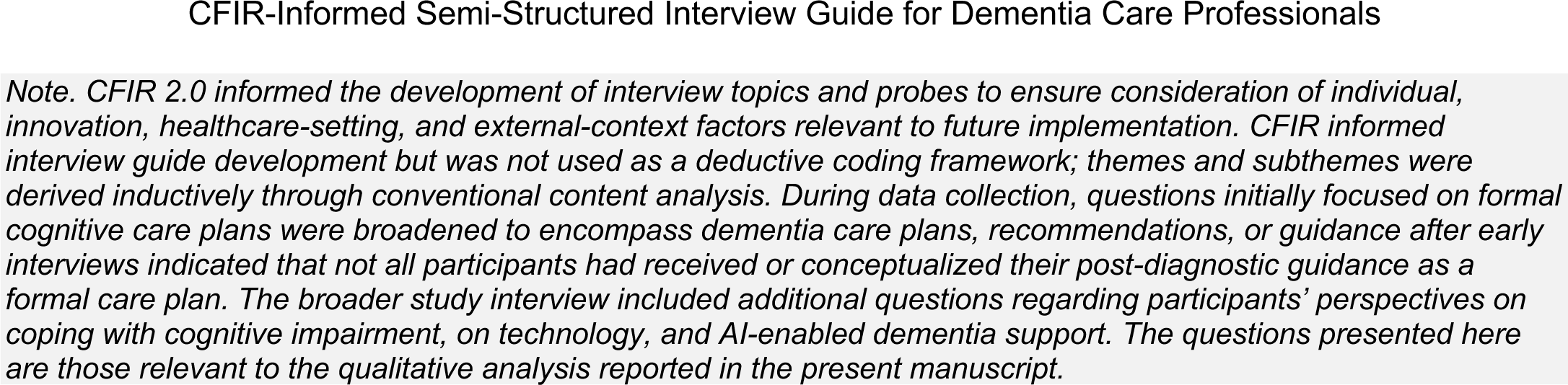

**Background**

*CFIR 2.0 domains informing this section: Individuals; Inner Setting*

1. Can you tell me about **your role** and your experience providing **care for patients** living with MCI or dementia?
2. Do you involve or provide any cognitive care planning, recommendations, or guidance to them?

Challenges and Questions Related to Dementia Care Plans, Recommendations, or Guidance

*CFIR 2.0 domains informing this section: Individuals; Outer Setting*

1. What are the most **<u>common challenges patients and care partners face</u>** when **<u>implementing</u>** cognitive care recommendations at home? (Open-ended)

a. What are the most common questions patients or care partners ask outside of scheduled appointments?
b. In your experience, what types of information do patients and care partners struggle to understand or remember?
2. Thinking about dementia care plans, recommendations, or guidance, what challenges or types of support do you think patients and care partners might need to use them in their daily lives?

**Probe …into these various Domains/Topics in CCP: What about in…

● Advance care planning and palliative care needs?
● Community resources including respite services, adult day care programs, or support groups?
● Care partner knowledge, care partner needs, social supports, & cg willingness to take on care partner tasks
● Safety (home – security, cooking safety, weapons removed, clutter, driving – vehicle)
● Neuropsychiatric and behavioral symptoms (depression, delusion, hallucination, apathy, etc.)
● Medication questions?
● Functional health (ADLs (Activities of Daily Life), Independent Activities of Daily LIving (IADLs))?
● Medical decision making?
● Cognitive (completing tasks, memory, reminders, etc.)

1. What are the most frequent questions patients and care partners with mild cognitive impairment or mild dementia and care partners ask regarding dementia care plans, recommendations, or guidance? (Open-ended)

What are the most frequently asked questions regarding…..[Considering the various Domains/Topics in CCP]…

● Advanced care planning and palliative care needs?
● Community resources including respite services, adult day care programs, support groups?
● Care partner knowledge, care partner needs, social supports, & cg willingness to take on care partner tasks?
● Safety (home – security, cooking safety, weapons removed, clutter, driving – vehicle)?
● Neuropsychiatric and behavioral symptoms (depression, delusion, hallucination, apathy, etc.)?
● Medication questions?
● Functional health (ADLs, IADLs)?
● Medical decision-making?
● Cognitive tasks (completing tasks, memory, reminders, etc.)?
● Communication

**Adoption and Support**

*CFIR 2.0 domains informing this section: Individuals; Innovation; Inner Setting; Outer Setting*

1. In your experience, what are the biggest barriers preventing successful implementation of dementia care plans, recommendations, or guidance? (Open-ended)

● what about any financial barriers?
● what about any social barriers?
● any cultural barriers? (i.e., gender, language/communications, traditions/customs, social norms, beliefs/values, dietary, lifestyles, habits, race)
● any logistical barriers? (operations? workflow in practice? people involved, time, etc)
2. In your experience, what facilitates implementation of dementia care plans, recommendations, or guidance? (Open-ended)

● what about any financial facilitators?
● what about any social facilitators?
● any cultural facilitators? (i.e., gender, language/communications, traditions/customs, social norms, beliefs/values, dietary, lifestyles, habits, race)
● any logistical facilitators?
● what additional support?
● what additional resources?
3. What have you found to be the most helpful for supporting dementia dyads in using dementia care plans, recommendations, or guidance? (Open-ended)

a. Are there specific community resources that are effective?
b. Are there any specific tools that are effective?
c. Are there any specific interventions that are effective? (examples? access, lifestyle change, support group, trainings, etc.)
4. What advice or recommendations do you most often provide to care partners related to dementia care plans, recommendations, or guidance? (Open-ended)
5. Are there specific concerns or misconceptions about dementia care plans, recommendations, or guidance that you often hear from care partners or patients? (Open-ended)
6. If you could improve one aspect of dementia care plans, recommendations, or guidance for dementia dyads, what would it be? (Open-ended)
7. Do you have any other comments or suggestions regarding dementia care plans, recommendations, or guidance that you would like to share? (Open-ended)

CFIR-Informed Semi-Structured Interview Guide for Dementia Dyads

*Note. CFIR 2.0 informed the development of interview topics and probes to ensure consideration of individual, innovation, healthcare-setting, and external-context factors relevant to future implementation. CFIR informed interview guide development but was not used as a deductive coding framework; themes and subthemes were derived inductively through conventional content analysis. During data collection, questions initially focused on formal cognitive care plans were broadened to encompass dementia care plans, recommendations, or guidance after early interviews indicated that not all participants had received or conceptualized their post-diagnostic guidance as a formal care plan. The broader study interview included additional questions regarding participants’ perspectives on coping with cognitive impairment, on technology, and AI-enabled dementia support. The questions presented here are those relevant to the qualitative analysis reported in the present manuscript*.

**General Dementia Care Understanding**

*CFIR 2.0 domains informing this section: Innovation; Individuals*

1. Can you describe your most recent cognitive care plan, recommendations, or guidance?

● When did you receive it?
● What does it include?
2. What does the care plan, recommendations, or guidance include or focus on? (e.g., medication, safety, memory, routines, daily tasks)
3. Do you use the care plan, recommendations, or guidance in daily life?

● If yes, how often do you follow it?
● If no, why not?
4. Was anything in the care plan, recommendations, or guidance new or surprising to you?

● If yes, what stood out?
5. Do you find the care plan, recommendations, or guidance helpful? Why or why not?

● If yes, what parts have been most helpful?
● If no, why not?
6. Was any part of the care plan, recommendations, or guidance confusing or hard to understand?

● If yes, which parts and what makes them unclear?

Did you ever receive information that felt unclear, overwhelming, or even contradictory?

What specific strategies, tools, or routines ended up working best for you?

1. Does the care plan, recommendations, or guidance reflect your or your loved one’s unique needs, values, or preferences?

● If yes, in what ways?
● If no, why not?

**Implementation Experience**

*CFIR 2.0 domains informing this section: Innovation; Individuals*

1. What challenges have you faced (or expect to face) in using the care plan, recommendations, or guidance day to day?

● Challenges for the patient
● Challenges for the caregiver
2. What parts of the care plan, recommendations, or guidance have been easy to implement?
3. Have there been any specific activities or recommendations that are difficult to implement?

● If yes, which ones and how are they difficult?
4. Have there been times when your loved one resisted specific aspects of the care plan, recommendations, or guidance?

● If yes, which ones and how did you handle those situations?
5. Have there been times when you felt unsure about what to do in a particular caregiving situation?

● If yes, can you describe one? What did you do? Who do you usually look to for help?

**Support & Resource Needs**

*CFIR 2.0 domains informing this section: Individuals; Inner Setting; Outer Setting*

1. Have you received any guidance or training on how to follow the care plan, recommendations, or guidance?

● If yes, who provided it and was it helpful?
● If not, would that have been helpful?
2. What kind of support would help you follow the care plan, recommendations, or guidance more easily? Why?
3. Do you feel supported by healthcare professionals in managing the care plan, recommendations, or guidance?

● If yes, what kind of support are you getting?
● If not, what kind of support do you wish you had?
4. Have you used any community services or resources for support?

● If yes, what did you use and how helpful was it?
● If not, has anything made it hard to access those resources?
5. What other types of support or services do you wish were available to help you?

**Key Care Challenges & Support Areas**

*CFIR 2.0 domains informing this section: Individuals; Outer Setting*

1. Are there areas in your care plan, recommendations, or guidance where you’d like more information or support, either in the past or currently?

● Safety
● Advanced care planning (e.g., living will, power of attorney)
● Medications
● Behaviors (neuropsychiatric symptoms)
● Communication
● Functional health (ADLs, IADLs)
● Other health conditions
● Medical decisions
● Cognitive function

**Safety**

● Do you have any safety concerns at home?

○ If so, what are they?
● Have you talked about stopping driving?

○ Have you made a plan to stop driving?
○ Any difficulties with that?

**Advance Care Planning**

● Do you have any questions about

○ Planning for future care preferences?
○ Choosing someone to make decisions for you about money or health?
○ Have you spoken with your loved one about this?

▪ If yes, how did it go?

▪ If not, what’s made it difficult?

▪ Have you prepared any documents such as a living will, Power of Attorney, or any documents for decisions about money and health?

**Medication Management**

● How do you currently manage medications? Are there any difficulties?

○ Have you ever missed or had difficulty keeping track of medications? If so, what contributed to that?
○ Is the patient currently participating in any clinical trials or using any investigational treatments? If so, what are your experiences?
○ What support or tools do you feel would help improve medication management, if needed?

**Behavioral Symptoms**

● Have you noticed any behavioral changes?

○ Any agitation, aggression, or withdrawal?
○ What symptoms have been the most difficult to handle?
○ How do you manage them? Has it helped?
● Do you feel you have enough guidance/resources to address behavioral symptoms?

○ If not, what would be helpful to add?

**Communication**

● Have you had communication challenges?

○ Please describe them.
● How have you addressed them?

○ What has helped?

**Functional Health (ADLs/IADLs)**

● Any difficulties with basic daily tasks, such as bathing, dressing, toileting, meal prep, etc.?

○ Please describe them.
○ How are you managing them?
● Any difficulties with daily tasks, such as shopping, cooking, housekeeping, using transportation, managing finances, etc.?

○ Please describe them.
○ How are you managing them?

**Physical Health & Comorbidities**

● How do you manage other chronic health conditions?

○ Do you have any questions about how to manage these alongside dementia care?
● Any challenges coordinating medical care for multiple conditions?
● How do nutrition and exercise factor into your care, any challenges?

**Medical Decision-Making**

● How do you make healthcare decisions together?

○ Have there been any particularly difficult medical decisions? Please describe.
○ What support might help?

**Cognitive Functioning**

● What cognitive symptoms (e.g., memory loss, language difficulties, reasoning challenges, issues with completing tasks, etc.) are most noticeable?

○ How do they affect daily life?
○ How are you addressing them?

Are there any other challenges you’re facing right now that we haven’t talked about yet?

**Reflections & Recommendations**

*CFIR 2.0 domains informing this section: Individuals; Innovation; Inner Setting; Outer Setting*

**Reflections**

1. What advice would you give to other families who are facing similar challenges in implementing cognitive care plans, recommendations, or guidance?
2. If you could change one thing about the support or guidance you’ve received for dementia care at home, what would it be and why?
3. What has helped you the most in navigating this journey — whether emotionally, practically, or spiritually?

Looking back now, is there anything you understand differently about the caregiving experience?

**Recommendations**

1. What recommendations do you have for healthcare providers to improve dementia care planning, recommendations, or guidance for families like yours?
2. Do you think more education or guidance related to dementia care planning and recommendations should be provided to patients and caregivers? Why or why not?

## References

1. ADI - Dementia statistics. (n.d.). Retrieved August 26, 2026, from https://www.alzint.org/about/dementia-facts-figures/dementia-statistics/

2. Bamford, C., Wheatley, A., Brunskill, G., Booi, L., Allan, L., Banerjee, S., Dening, K. H., Manthorpe, J., Robinson, L., & Team, on behalf of the P. study. (2021). Key components of post-diagnostic support for people with dementia and their carers: A qualitative study. PLOS ONE, 16(12), e0260506. 10.1371/journal.pone.0260506

3. Bastholm-Rahmner, P., Schmidt-Mende, K., Modig, K., & Bergqvist, M. (2025). Living well with dementia: A qualitative interview study on family caregivers’ call for more person- and family-centered dementia support. BMC Geriatrics, 25(1), 758. 10.1186/s12877-025-06429-z

4. Berghout, M., Waller, A., Lachapelle, N., Noble, N., Nair, B. K., & Sanson-Fisher, R. (2024). Preferred type, timing and format of dementia information: A cross-sectional survey of carers of people living with dementia. Australasian Journal on Ageing, 43(1), 131–139. 10.1111/ajag.13251

5. Bergqvist, M., Bastholm-Rahmner, P., Modig, K., & Schmidt-Mende, K. (2026). Challenges and needs in dementia care: People with dementia and family caregivers’ experiences from diagnosis to follow-up in Swedish primary care: a qualitative study. Scandinavian Journal of Primary Health Care, 44(1), 1–14. 10.1080/02813432.2025.2529414

6. Bressan, V., Visintini, C., & Palese, A. (2020). What do family caregivers of people with dementia need? A mixed-method systematic review. Health & Social Care in the Community, 28(6), 1942–1960. 10.1111/hsc.13048

7. Britt, K. C., Xiao, X., Sun, K., Ge, S., Buck, H., & Huang, B. (2025). A qualitative study with primary care providers: Barriers and facilitators in cognitive care planning. Geriatric Nursing (New York, N.Y.), 67, 103726. 10.1016/j.gerinurse.2025.103726

8. Caprioli, T., Mason, S., Tetlow, H., Reilly, S., & Giebel, C. (2023). Exploring the views and the use of information and communication technologies to access post-diagnostic support by people living with dementia and unpaid carers: A systematic review. Aging & Mental Health, 27(12), 2329–2345. 10.1080/13607863.2023.2196246

9. Cipriani, G., Danti, S., Picchi, L., Nuti, A., & Fiorino, M. D. (2020). Daily functioning and dementia. Dementia & Neuropsychologia, 14(2), 93–102. 10.1590/1980-57642020dn14-020001

10. Damschroder, L. J., Reardon, C. M., Widerquist, M. A. O., & Lowery, J. (2022). The updated Consolidated Framework for Implementation Research based on user feedback. Implementation Science: IS, 17(1), 75. 10.1186/s13012-022-01245-0

11. Efthymiou, A., Papastavrou, E., Middleton, N., Markatou, A., & Sakka, P. (2020). How Caregivers of People With Dementia Search for Dementia-Specific Information on the Internet: Survey Study. JMIR Aging, 3(1), e15480. 10.2196/15480

12. Fowler, N. R., Partrick, K. A., Taylor, J., Hornbecker, M., Kelleher, K., Boustani, M., Cummings, J. L., MacLeod, T., Mielke, M. M., Brosch, J. R., Lee, J., Shobin, E., Galvin, J. E., Fillit, H., Udeh-Momoh, C., & Willis, D. R. (2025). Implementing early detection of cognitive impairment in primary care to improve care for older adults. Journal of Internal Medicine, 298(1), 31–45. 10.1111/joim.20098

13. Frederiksen, K. S., Arus, X. M., Zetterberg, H., Gauthier, S., Boada, M., Pytel, V., Hahn-Pedersen, J., Solís Tarazona, L. R., & Mattke, S. (2024). Focusing on earlier diagnosis of Alzheimer’s disease. Future Neurology, 19(1), 2337452. 10.2217/fnl-2023-0024

14. Ge, S., Xiao, X., Huang, B., & Britt, K. C. (2025). Exploring neurologists’ perspectives: Barriers and facilitators in implementing cognitive care planning. The Public Policy and Aging Report, 35(3), 70–75. 10.1093/ppar/praf014

15. Grand, J. H., Caspar, S., & MacDonald, S. W. (2011). Clinical features and multidisciplinary approaches to dementia care. Journal of Multidisciplinary Healthcare, 4, 125–147. 10.2147/JMDH.S17773

16. Hennink, M. M., Kaiser, B. N., & Marconi, V. C. (2017). Code Saturation Versus Meaning Saturation: How Many Interviews Are Enough? Qualitative Health Research, 27(4), 591–608. 10.1177/1049732316665344

17. Hsieh, H.-F., & Shannon, S. E. (2005). Three approaches to qualitative content analysis. Qualitative Health Research, 15(9), 1277–1288. 10.1177/1049732305276687

18. Kallmyer, B. A., Bass, D., Baumgart, M., Callahan, C. M., Dulaney, S., Evertson, L. C., Fazio, S., Judge, K. S., & Samus, Q. (2023). Dementia care navigation: Building toward a common definition, key principles, and outcomes. Alzheimer’s & Dementia, 9(3), e12408. 10.1002/trc2.12408

19. Kern, L. M., Riffin, C., Phongtankuel, V., Aucapina, J. E., Banerjee, S., Ringel, J. B., Tobin, J. N., Fisseha, S., Meiri, H., Bell, S. K., & Casale, P. N. (2024). Gaps in the coordination of care for people living with dementia. Journal of the American Geriatrics Society, 72(10), 3119–3128. 10.1111/jgs.19105

20. Kim, H., Sefcik, J. S., & Bradway, C. (2017). Characteristics of Qualitative Descriptive Studies: A Systematic Review. Research in Nursing & Health, 40(1), 23–42. 10.1002/nur.21768

21. Kokorelias, K. M., Shiers-Hanley, J. E., Li, Z., & Hitzig, S. L. (2023). A Systematic Review on Navigation Programs for Persons Living With Dementia and Their Caregivers. The Gerontologist, 63(8), 1341–1350. 10.1093/geront/gnac054

22. Lincoln, Y. S., & Guba, E. G. (1985). Naturalistic Inquiry. SAGE.

23. Maddali, H. T., Dixon, E., Pradhan, A., & Lazar, A. (2022). Investigating the Potential of Artificial Intelligence Powered Interfaces to Support Different Types of Memory for People with Dementia. Extended Abstracts on Human Factors in Computing Systems. CHI Conference, 226(226), 10.1145/3491101.3519858. 10.1145/3491101.3519858

24. Mueller, A., Thao, L., Condon, O., Liebzeit, D., & Fields, B. (2022). A Systematic Review of the Needs of Dementia Caregivers Across Care Settings. Home Health Care Management & Practice, 34(2), 133–147. 10.1177/10848223211056928

25. Ng, K., Martin-Khan, M., Farrow, M., Beattie, E., & Pachana, N. (2016). The Implications of the Timing of Diagnosis of Dementia on the Dementia Caregiver. 5(4), 143–154. 10.4236/aad.2016.54010

26. Shafir, A., Ritchie, C. S., Garrett, S. B., Sideman, A. B., Naasan, G., Merrilees, J., Widera, E., Flint, L., & Harrison, K. L. (2022). “Captive by the uncertainty” – Experiences with anticipatory guidance for people living with dementia and their caregivers at a specialty dementia clinic. Journal of Alzheimer’s Disease : JAD, 86(2), 787–800. 10.3233/JAD-215203

27. Shi, J. M., Yoo, D. W., Wang, K., Rodriguez, V. J., Karkar, R., & Saha, K. (2026). Mapping Caregiver Needs to AI Chatbot Design: Strengths and Gaps in Mental Health Support for Alzheimer’s and Dementia Caregivers. ACM Transactions on Computing for Healthcare. 10.1145/3803549

28. Soong, A., Au, S. T., Kyaw, B. M., Theng, Y. L., & Tudor Car, L. (2020). Information needs and information seeking behaviour of people with dementia and their non-professional caregivers: A scoping review. BMC Geriatrics, 20(1), 61. 10.1186/s12877-020-1454-y

29. Sorrentino, M., Mercogliano, M., Fiorilla, C., Stilo, I., Esposito, F., Moccia, M., Lavorgna, L., Affinito, G., Salvatore, E., Sormani, M. P., Odone, A., Majeed, A., Rubba, F., Triassi, M., & Palladino, R. (2025). The needs and unmet needs for people living with dementia, caregivers and care workers in dementia health care systems: A systematic review. Frontiers in Public Health, 13, 1605993. 10.3389/fpubh.2025.1605993

30. Teles, S., Paúl, C., Costa-Santos, C., & Ferreira, A. (2022). Use of dementia and caregiving-related internet resources by informal caregivers: A cross-sectional study. Frontiers in Medicine, 9, 978635. 10.3389/fmed.2022.978635

31. Tong, A., Sainsbury, P., & Craig, J. (2007). Consolidated criteria for reporting qualitative research (COREQ): A 32-item checklist for interviews and focus groups. International Journal for Quality in Health Care: Journal of the International Society for Quality in Health Care, 19(6), 349–357. 10.1093/intqhc/mzm042

32. van der Roest, H. G., Meiland, F. J. M., Comijs, H. C., Derksen, E., Jansen, A. P. D., van Hout, H. P. J., Jonker, C., & Dröes, R.-M. (2009). What do community-dwelling people with dementia need? A survey of those who are known to care and welfare services. International Psychogeriatrics, 21(5), 949–965. 10.1017/S1041610209990147

33. Wasilewski, M. B., Stinson, J. N., & Cameron, J. I. (2017). Web-based health interventions for family caregivers of elderly individuals: A Scoping Review. International Journal of Medical Informatics, 103, 109–138. 10.1016/j.ijmedinf.2017.04.009

34. Yang, K. L., Kelble, L., Felten, K., Carlsson, C. M., & Clark, L. R. (2025). Memory screening in the community: Facilitating earlier dementia diagnosis and care—Preliminary data. Journal of the American Geriatrics Society, 73(4), 1227–1236. AgeLine (184494052). 10.1111/jgs.19302

35. Zuber, M., Borate, S. N., Tan, C. J., & Li, N. (2026). Health Care-Related Tasks of Informal Dementia Caregivers in the U.S.: A Systematic Review with Narrative Synthesis. *Inquiry: A Journal of Medical Care Organization*, Provision and Financing, 63, 469580261438330. 10.1177/00469580261438330

